# Optimal Allocation of COVID-19 Test Kits Among Accredited Testing Centers in the Philippines

**DOI:** 10.1101/2020.04.14.20065201

**Authors:** Christian Alvin H. Buhat, Jessa Camille C. Duero, Edd Francis O. Felix, Jomar F. Rabajante, Jonathan B. Mamplata

## Abstract

Testing is crucial for early detection, isolation, and treatment of coronavirus disease (COVID-19)-infected individuals. However, in resource-constrained countries such as the Philippines, test kits have limited availability. As of 12 April 2020, there are 11 testing centers in the country that have been accredited by the Department of Health (DOH) to conduct testing. In this paper, we determine the optimal percentage allocation of COVID-19 test kits among accredited testing centers in the Philippines that gives an equitable chance to all infected individuals to be tested. Heterogeneity in testing accessibility, population density of municipalities, and the capacity of testing facilities are included in the model. Our results showed that the range of optimal allocation per testing center are: Research Institute for Tropical Medicine (4.17% −6.34%), San Lazaro Hospital (14.65% −24.03%), University of the Philippines-National Institutes of Health (16.25% −44.80%), Lung Center of the Philippines (15.8% −26.40%), Baguio General Hospital Medical Center (0.58% −0.76%), The Medical City, Pasig City (5.96% −25.51%), St. Luke’s Medical Center, Quezon City (1.09% −6.70%), Bicol Public Health Laboratory (0.06% −0.08%), Western Visayas Medical Center (0.71% −4.52%), Vicente Sotto Memorial Medical Center (1.02% −2.61%), and Southern Philippines Medical Center (≈0.01%). If there will be changes in the number of testing centers, our model can still be used to modify the test kit allocation. Our results can serve as a guide to the authorities in distributing the COVID-19 test kits. These can also be used to determine the capacity of testing centers and the effects of increasing its number. The model can also be used for proposing additional number and location of new testing centers.

## 1. Introduction

Affecting more than 170 countries, the coronavirus disease (COVID-19) has been declared by the World Health Organization (WHO) as a pandemic [16]. As of 12 April 2020, there are almost 1.7 million confirmed COVID-19 cases worldwide and about 4,500 cases in the Philippines [6, 9]. However, this number may not be an accurate representation of the total number of infected individuals in the Philippines because of its selective testing programs due to the limited availability of test kits [10]. Only those who are considered “patients under investigation” (PUI) are qualified to be tested. These are persons with severe symptoms such as fever and/or respiratory syndrome, history of travel in the past 14 days to countries with local transmission and history of exposure with a confirmed case [8].

The Department of Health (DOH) of the Philippines has already accredited 11 laboratories and hospitals (as of 12 April 2020) to conduct COVID-19 testing in the country. These centers have conducted a total of 38,640 tests (as of 11 April 2020) [12, 9]. The testing centers are Research Institute for Tropical Medicine (RITM), San Lazaro Hospital, University of the Philippines-National Institutes of Health (UP-NIH), Lung Center of the Philippines, Baguio General Hospital Medical Center (BGHMC), The Medical City, Pasig City (TMC), St. Luke’s Medical Center, Quezon City, Bicol Public Health Laboratory (BPHL), Western Visayas Medical Center (WVMC), Vicente Sotto Memorial Medical Center (VSMMC), and Southern Philippines Medical Center (SPMC). With these, and several other laboratories awaiting accreditation, the Philippine government plans to start a massive testing program for COVID-19 on 14 April 2020 to fully assess the public health situation. Conducting mass testing will not only help in tracing the asymptomatic carriers who need isolation, it will also protect the healthcare workers who are exposed to this contagious disease [10]. Even though DOH will increase its capacity for testing, resources such as testing kits are still insufficient to efficiently test the whole susceptible population in the Philippines [5, 13]. Thus, it is necessary to find an optimal strategy of distributing the available test kits among the accredited testing centers such that each individual estimated to be infected has an equitable chance of being tested.

In this paper, we use a Non-Linear Programming (NLP) model on designing equitable resource allocation on resource-constrained countries to address this issue. The mathematical model includes heterogeneity in testing accessibility, and the data are population-density based. The equity objective function of the model provides an equal assessment of the distribution of the limited test kits among the testing centers. We apply the model to the population density-adjusted number of COVID-19 infected individuals projection in the Philippines. We let the basic reproduction number (average number of direct secondary cases generated from a contagious person in a fully susceptible community) of COVID-19 in the country to be *R*_0_ = 2.5 to estimate the infected number of individuals that should be tested [4, 11]. We also consider the density-based population of each municipalities in the country as well as the current testing centers accredited by the DOH.

The remaining parts of the paper is organized as follows: in Section 2, we discuss the preliminaries of the model: estimating the number of infected individuals, accessibility of testing kits and a model for the distribution of testing kits. We also develop the constraints which is then utilized in the NLP model formulated. In Section 3, we find the coordinates for the communities and determine the different values for the dispersal length scale parameter *k*. These values are used to determine the optimal allocation of testing kits distribution. Section 4 presents some conclusions and remarks. The last section provides the supplementary materials needed for the paper.

## 2. Model Formulation

### 2.1. Preliminaries

In this model, we consider the municipalities and cities in the Philippines to be the community *i* and estimate the number of infected individual in each community. We assume that these individuals will go to the nearest accredited testing center *j*.

### 2.1.1. Estimating the number of infected individuals

We consider a forecasted population of each communities in the Philippines for the year 2019 based on the 2015 Philippine Statistics Authority (PSA) Census [1]. Each population of the community is then multiplied to 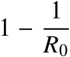with *R*_*0*_ = 2.5, this will give us the approximate cumulative number of infected individuals during the whole epidemic period per community on a do-nothing situation in a well-mixed system [14]. To relax the well-mixing property of the model, we then consider the population density of each community, and factor in a 1-meter additional distance (or 4*m*^2^ personal safe space) as recommended by the World Health Organization to observe social distancing. Thus, we compute for the projected total number of infected individuals in community *i, I*_*i*_, as

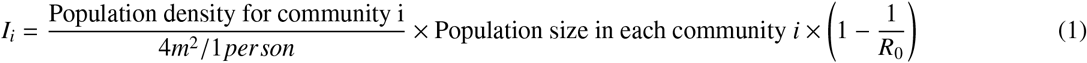

where population density is number of people per 1 million square meter.

#### 2.1.2. Computing for the Testing Accessibility

We assume that an individual tends to get tested in the nearest testing center *j*. The lesser the distance between the testing center and the person’s community, the greater the probability of the person going there. Let *d*_*i j*_ be the distance between community *i* and testing center *j* expressed as

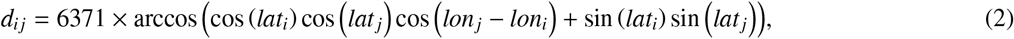

where *lat*_*i*_ and *lon*_*i*_ are the GPS location of community *i*, and *lat* _*j*_ and *lon*_*j*_ are are the GPS location of testing center *j*, all in radians measure. Using *d*_*i j*_, we then compute for the “effective demand” *f* (*d*_*i j*_)*I*_*i*_ or the number of infected individuals in community *i* that will go to testing center *j* for testing. We define *f* as a function that puts ‘weight’ on the testing accessibility of an individual to a testing center based on the distance *d*_*i j*_. To model the testing accessibility, we use a Gaussian model which is used to describe the distribution of the COVID-19 infections [15]. We have 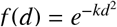, where *k* is a dispersal length scale parameter that can provide further information about outbreak dynamics and potential for superspreading events [11]. We use Elliot’s estimation in solving for *k* which considers the mean *µ*, variance *σ*^2^ of the distribution, and the sample size *N* for number of communities involved [7]:

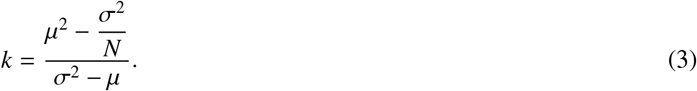

#### 2.1.3. Distribution of test kits

We follow the approach of Wilson and Blower [17] in determining the test kit distribution to each testing centers. First, we compute how many infected individuals will go to a certain testing center. We call this the demand *D*_*j*_ where

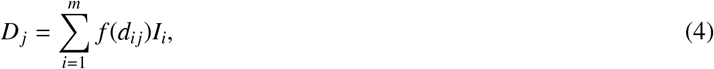

which is the sum of the effective demand of all communities to a testing center *j*. Let 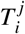 be the number of individuals that will get tested in each community *i* and it is given by

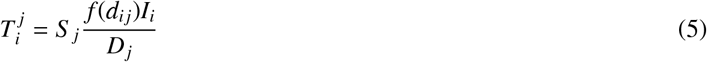

where *S*_j_ is the supply allocation on testing center *j* and 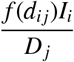 is the percent allocation of each testing center to each community *i*. Thus the total number of tested individuals in community *i, T*_*i*_, is given by

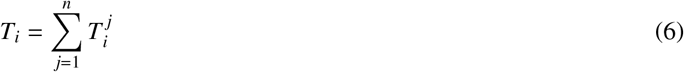

Each testing center has a maximum number of tests that they can perform per day, and thus we factor in the daily limit *L*_*j*_ of each testing center, and compute for the maximum number of testing days *M* as

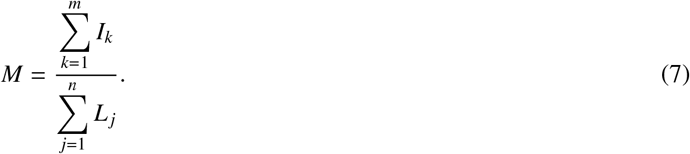

### 2.2. NLP Model

Nonlinear programming (NLP) is an optimization problem where the objective function is nonlinear and/or the feasible region is determined by nonlinear constraints [2]. Here, we use the NLP model based on Wilson-Blower [17], with COVID-19 data specific to the Philippines, to determine the optimal distribution of test kits among the testing centers. The model is described by the following:

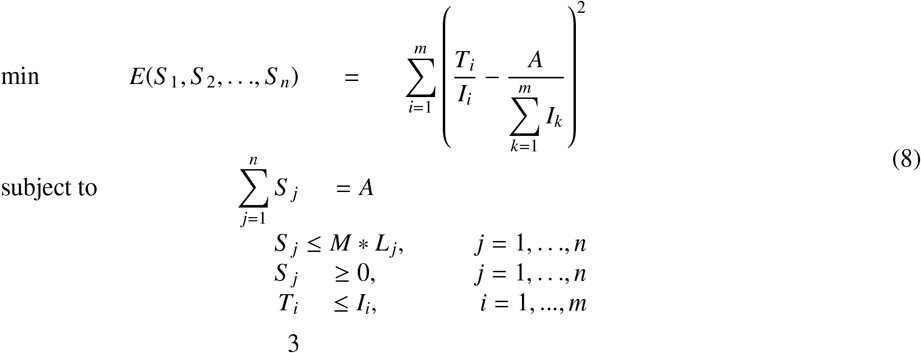

where *S* _*j*_ is the allocation of testing kits to testing center *j*, and *A* is the total number of testing kits to be allocated. Here, we use an equity objective function to ensure equal opportunities among all infected individuals in getting tested. We base the objective function to the Euclidean norm to provide a standard result in comparison with other norms [3]. We minimize the equity objective function *E*, with optimal allocations to each Testing Center (*S* _*j*_) such that: (i) all available testing kits will be used, (ii) daily limits of testing per testing center will be considered and will not exceed the maximum number of days for testing, (iii) there will be a non-negative supply of testing kits per testing center, and (iv) the number of tested individuals in each community is less than the expected maximum number of infected individuals. Note that each individual will only be subjected to one test, which will serve as an initial testing. We solve this NLP model through simulations in R and explore scenarios such as adding additional testing centers to communities with *I*_*i*_ ≥ 5000.

## 3. Numerical illustrations

We determine the GPS coordinates of all the cities and municipalities in the Philippines, and of the 11 accredited testing centers, using Geocode by Awesome Table app in google spreadsheets. This app returns the latitude and longitude coordinates of the testing centers’ locations and the geographic centers of the cities and municipalities. We use these coordinates in the computations except for two: Datu Hoffer Ampatuan and Shariff Sardona Mustapha, Maguindanao. Geocode app cannot locate Shariff Sardona Mustapha, and it returned incorrect coordinates for Datu Hoffer Ampatuan. We use the coordinates of one barangay from each municipality instead. In addition, we compute for various dispersal length scale parameters *k* shown in (3) using the mean, *µ*, and variance, *σ*^2^, of distributions of each density structures in the country : (i) population density-based distribution of infected individuals in the whole Philippines which resulted in *k* = 0.0849, (ii) distribution of infected individuals in Metro Manila and this results in *k* = 0.4136, and (iii) population density-based distribution of infected individuals in Metro Manila that results in *k* = 1.5614, the highest density in the country.

We utilize the 11 accredited testing centers to determine the optimal distribution of testing kits. Shown in Table 1 is the proportion of testing kits to be distributed among the 11 testing centers using different values for *k*. We can observe that UP-NIH will receive the biggest proportion, ranging from 16.25% to 44.80%, while Southern Philippines Medical Center will receive the smallest proportion which is 0.01%.

**Table 1:**
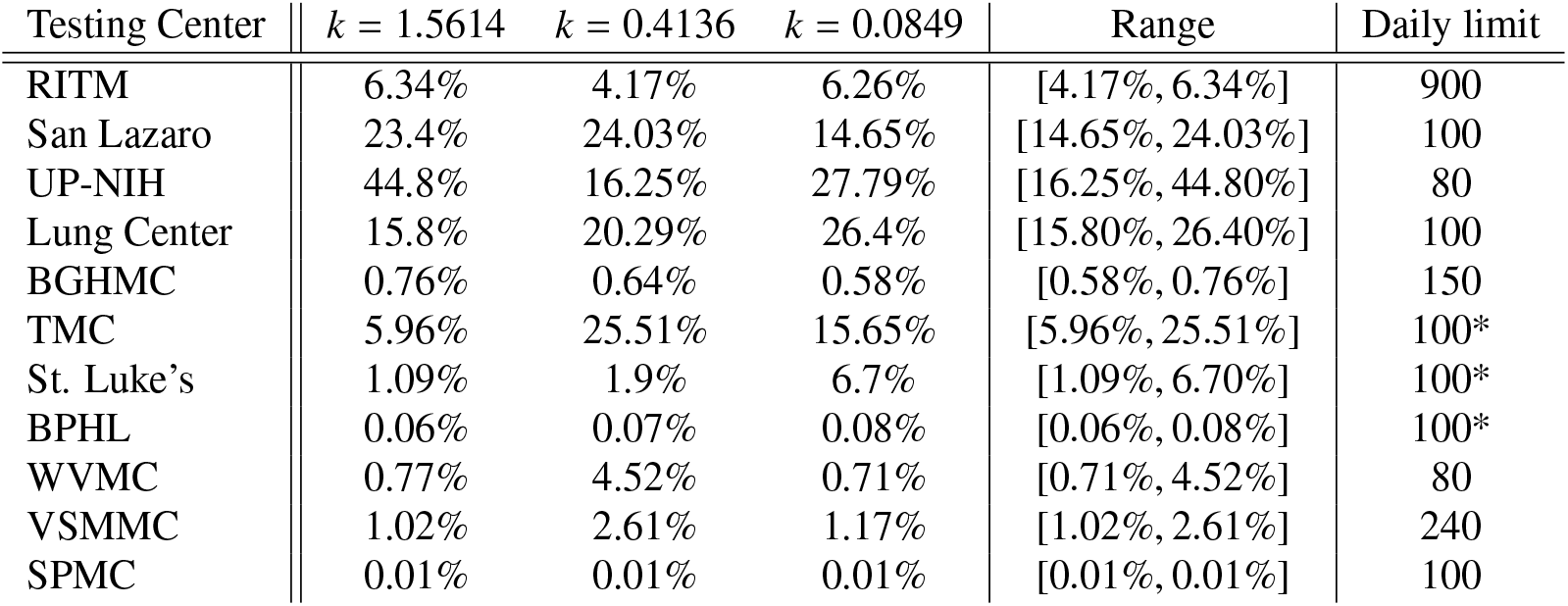
Proportion of test kits to be distributed in the different testing centers for *k* = 1.5614, *k* = 0.4136 and *k* = 0.0849 where * are assumed values for daily limit.

From the results, we notice that San Lazaro Hospital, UP-NIH and Lung Center of the Philippines got the majority of the proportion, across all scenarios. If mass testing will push through, these three hospitals will be overworked and will not be able to deliver timely results on the tests. To address this, we propose to add more testing centers. We identify 10 communities with the highest number of infected individuals and no existing accredited testing center. We will treat these 10 communities as the location of the suggested additional testing centers. We then compute for the optimal distribution of testing kits to the 21 testing centers. The proportion of test kits allocated to each of UP-NIH, San Lazaro Hospital, Lung Center of the Philippines and Makati City is greater than 10%. We can say that this distribution poses concerns similar to that of only having 11 testing centers. To reduce the proportion of test kits allocated to each of these testing centers, we consider all communities with more than 5,000 infected individuals. We then identify 19 communities as locations of new testing centers in addition to the original 11 and compute for the optimal distribution of testing kits to the 30 testing centers.

In addition, we obtain various sets of percentage allocations from different values of *k* representing different population density structures. Each *k* value from testing centers located in NCR (most populated region) vary in the percentage allocation per *k* (RITM, San Lazaro, UP-NIH, Lung Center, TMC, and St. Luke’s). One reason is that these testing centers have common infected individuals that are approximately close to multiple centers. This gives these individuals the option to go to their preferred testing center. Meanwhile, testing centers outside NCR remain fairly constant in terms of distribution, regardless of the value of *k*, since the same portion of infected individuals will get tested there.

Shown in Figure 2 are the pie charts representing the optimal distribution of testing kits for different number of testing centers and different values of dispersal length parameter. It is evident that as we increase the number of testing centers from 11 to 30, there is a great decrease of proportions to be distributed on the currently existing testing centers. Newly introduced testing centers will get a substantial amount of testing kits and will serve infected individuals in their respective vicinity.

**Figure 1:**
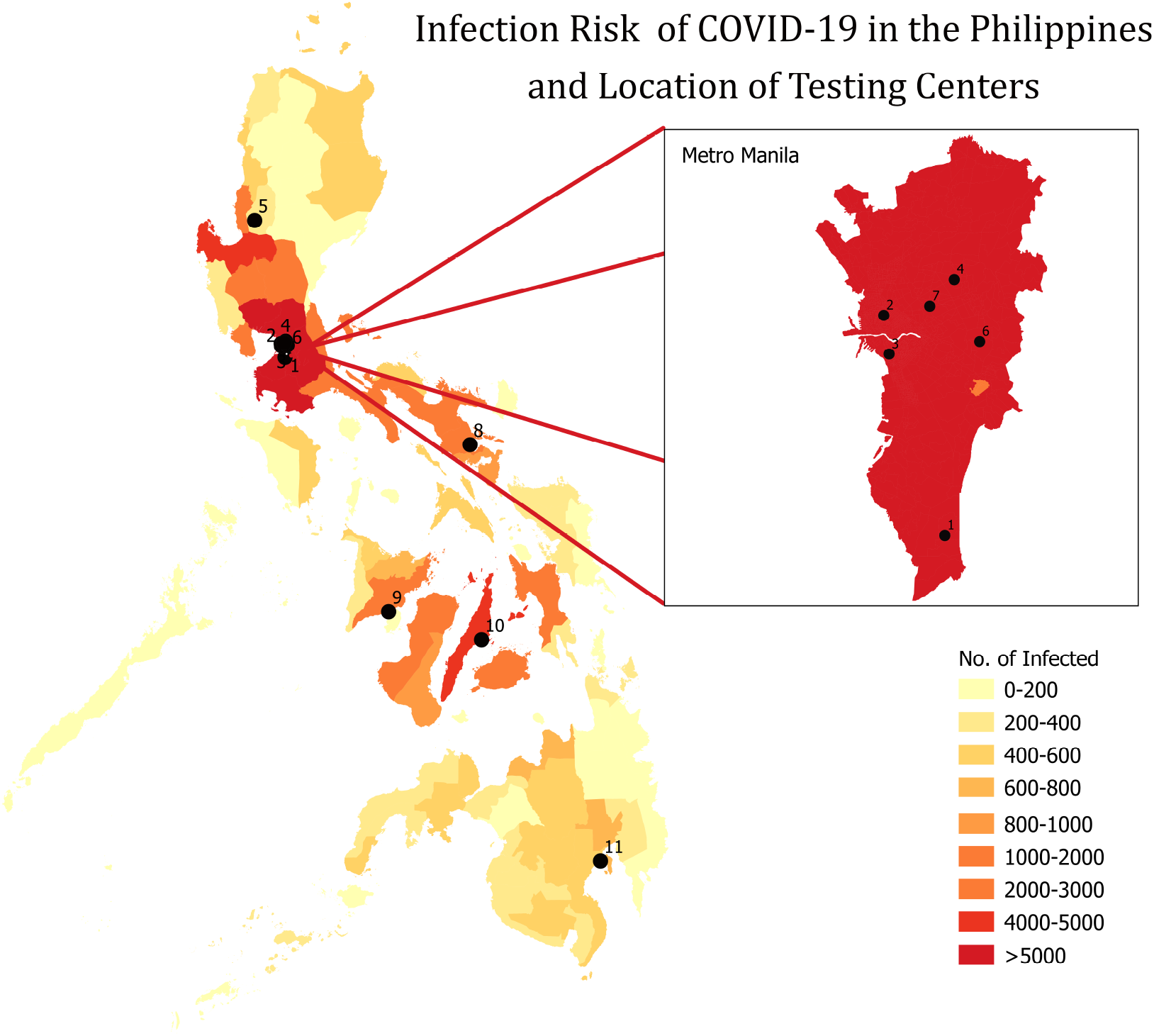
Heat map of infections and locations of testing centers accredited by the World Health Organization as of 12 April 2020. The following are the accredited testing centers and are shown on the map: 1. Research Institute for Tropical Medicine (RITM); 2. San Lazaro Hospital; 3. University of the Philippines - National Institutes of Health (UP-NIH); 4. Lung Center of the Philippines; 5. Baguio General Hospital and Medical Center (BGHMC); 6. The Medical City, Pasig City (TMC); 7. St. Luke’s Medical Center, Quezon City; 8. Bicol Public Health Laboratory (BPHL); 9. Western Visayas Medical Center (WVMC); 10. Vicente Sotto Memorial Medical Center (VSMMC); 11. Southern Philippines Medical Center (SPMC).

**Figure 2:**
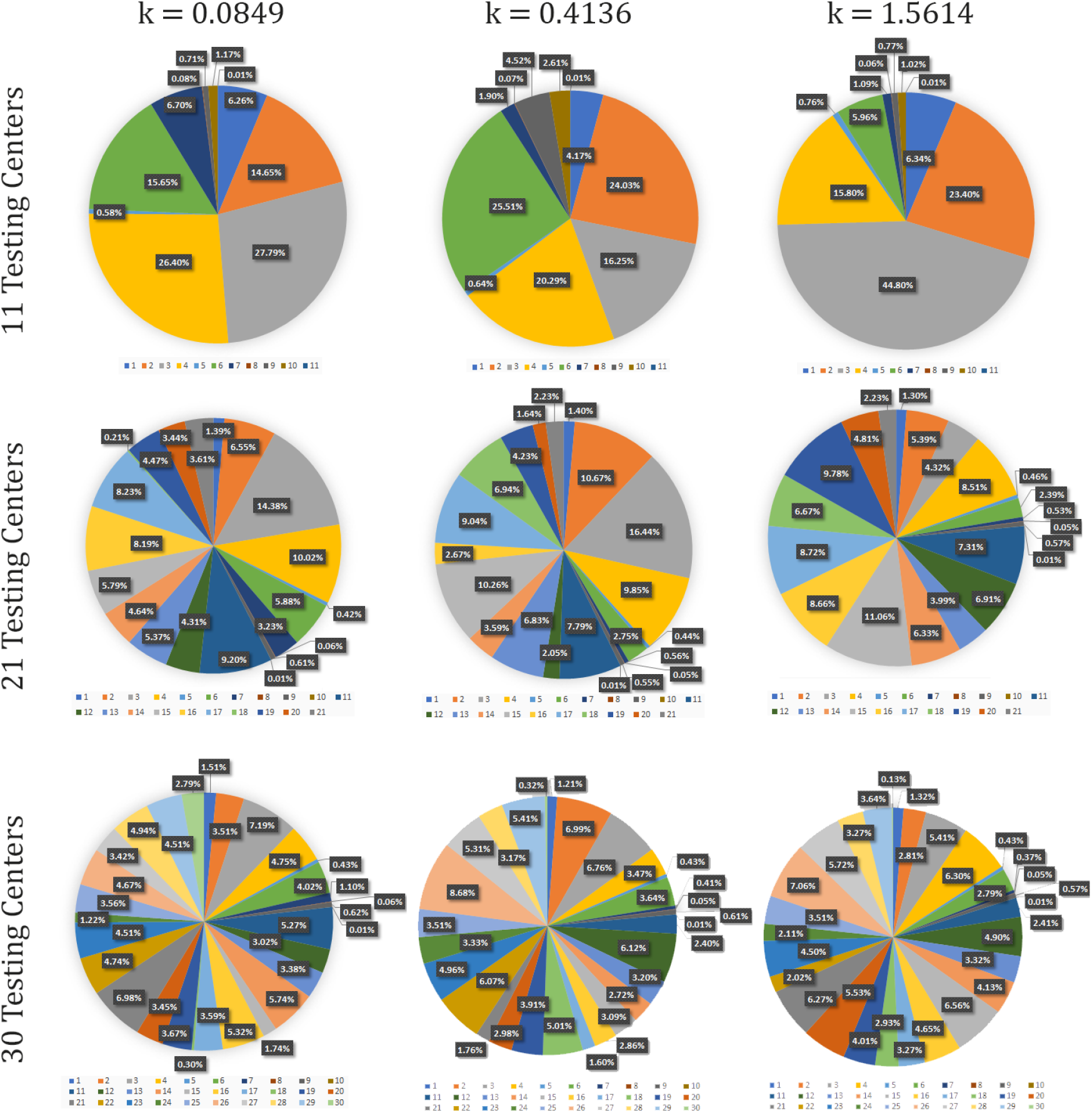
Proportion of testing kits for various number of testing centers against different values of *k*. These testing centers are located in: 1. Research Institute for Tropical Medicine (RITM); 2. San Lazaro Hospital; 3. University of the Philippines - National Institutes of Health; 4. Lung Center of the Philippines; 5. Baguio General Hospital and Medical Center; 6. The Medical City, Pasig City; 7. St. Luke’s Medical Center, Quezon City; 8. Bicol Public Health Laboratory; 9. Western Visayas Medical Center; 10. Vicente Sotto Memorial Medical Center; 11. Southern Philippines Medical Center; 12. Caloocan City; 13. Cavite; 14. City of Mandaluyong; 15. City of Makati; 16. Laguna; 17. Taguig City; 18. Pasay City; 19. Rizal; 20. Bulacan; 21. City of Las Piñas; 22. City of Paranãque; 23. City of Marikina; 24. City of Malabon; 25. City of Valenzuela; 26. City of Navotas; 27. Pangasinan; 28. Batangas; 29. Negros Occidental; 30. City of San Juan.

From the optimal solution computed using 30 testing centers, we can see that the maximum proportion of testing kits to be distributed is in Navotas City, which accounts for 8.68% of the total number of testing centers. This is achieved for *k* = 0.4136. Furthermore, we can infer that adding more testing centers will greatly reduce the work to be done by each testing centers. Shown in Figure 3 is the map with corresponding locations of the 30 testing centers.

**Figure 3:**
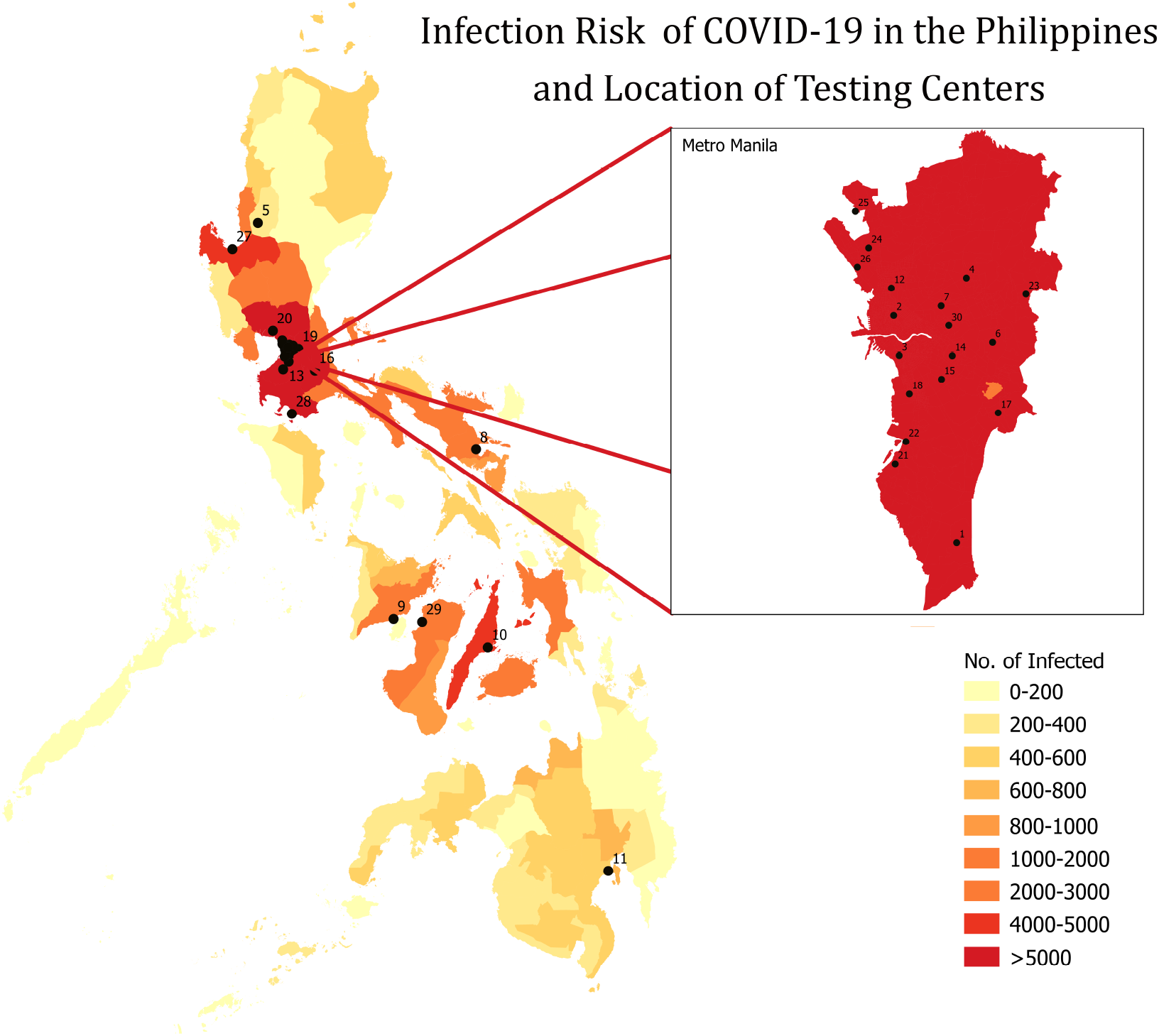
Locations of the 11 testing centers accredited by the World Health Organization as of 12 April 2020 and the 19 additional testing centers. The following are the testing centers and are shown in the map: 1. Research Institute for Tropical Medicine (RITM); 2. San Lazaro Hospital; 3. University of the Philippines - National Institutes of Health (UP-NIH); 4. Lung Center of the Philippines; 5. Baguio General Hospital and Medical Center (BGHMC); 6. The Medical City, Pasig City (TMC); 7. St. Luke’s Medical Center, Quezon City; 8. Bicol Public Health Laboratory (BPHL); 9. Western Visayas Medical Center (WVMC); 10. Vicente Sotto Memorial Medical Center (VSMMC); 11. Southern Philippines Medical Center (SPMC); 12. Caloocan City; 13. Cavite; 14. City of Mandaluyong; 15. City of Makati; 16. Laguna; 17. Taguig City; 18. Pasay City; 19. Rizal; 20. Bulacan; 21. City of Las Pinãs; 22. City of Paran ã que; 23. City of Marikina; 24. City of Malabon; 25. City of Valenzuela; 26. City of Navotas; 27. Pangasinan; 28. Batangas; 29. Negros Occidental; 30. City of San Juan.

## 4. Discussion

We determined an equitable and optimal allocation of COVID-19 testing kits among the DOH accredited testing centers in the Philippines. We applied Wilson-Blower’s Nonlinear Programming Model, which gave an equitable opportunity to each susceptible individual to be tested. The model involved COVID-19 specific conditions such as infected population, reproduction number of the disease (*R*_0_ = 2.5), distribution of infected individuals, and testing limits of each testing centers. We applied this to the municipality/city data of the Philippines and computed the percentage distribution on different dispersal length parameter *k* values, *k* = 0.0849, *k* = 0.4136, and *k* = 1.5614.

We ran computations using R on the model to determine the optimal allocation of testing kits to be distributed amongst accredited DOH testing centers and additional testing centers found in highly-infected communities in the country. Based on our results, here is the optimal range of distribution per testing center: RITM (4.17% −6.34%), San Lazaro (14.65% 24.03%), UP-NIH (16.25% −44.80%), Lung Center (15.8% −26.40%), BGHMC (0.58% −0.76%), TMC (5.96% −25.51%), St. Luke’s, QC (1.09% −6.70%), BPHL (0.06% −0.08%), WVMC (0.71% −4.52%), VSMMC (1.02% −2.61%), and SPMC (≈ 0.01%).

Note that if only 11 testing centers are to be used for testing COVID-19 patients, several testing centers will be overworked, especially if mass testing will push through. For instance, let us assume that 300,000 testing kits will be distributed. From the optimal distribution, UP-NIH will receive an average of almost 89,000 testing kits, across the different values we considered for the dispersal length. This implies that it needs more than 1,000 days to complete the testing for patients in UP-NIH which translates to more than two years to complete the testing. If we consider the proposed 30 testing centers, UP-NIH will receive almost 19,000 testing kits, and this translates to 245 days to complete the testing. This is a drastic decrease to the number of days needed to complete all testing from 11 testing centers to 30 testing centers. With these realizations, we observed that the greater the number of testing centers operating, the faster the completion of all testing of patients, and the more patients tested per day. We also recommend that these additional testing centers should be located in strategic places, that is, in the location with high density of infected individuals as shown in Figure 3.

The model is helpful to policy makers in making a decision on how to distribute available COVID-19 test kits among accredited testing centers. Authorities may apply this to their respective communities, if the public health system and situation follow the same assumptions and goal of the model. In addition, they may also assess the feasibility of the percentage allocation depending on their current situation. The model exhausts all available test kits but the complexity of test kit resource dynamics determines the limitation of our study. Further studies should also incorporate individuals needing multiple tests, effectiveness of testing kits, and transport time for testing kit delivery. In conclusion, a larger amount of test kits (not limited to initial testing, and to symptomatic patients) and more testing centers are ideal to increase the number of individuals tested to aid in “flattening” the epidemic curve.

## Data Availability

All data and program used are available online at: https://github.com/alvinizer/optimalallocationPH

## 5. Supplementary Files

The Microsoft Excel file containing the municipality/city data, GPS location, and population density-based number of infections, and R files used for the model simulation can be found online at: github.com/alvinizer/optimalallocationPH/.

## 6. Acknowledgment

JFR is supported by the Abdus Salam International Centre for Theoretical Physics Associateship Scheme.

## Appendix A. Optimal solution for 21 testing centers

**Table A.2:** Proportion of test kits to be distributed in 21 testing centers.

| Testing Center | $k = 1.5614$ | $k = 0.4136$ | $k = 0.0849$ | Range | Daily limit |
| --- | --- | --- | --- | --- | --- |
| RITM | 1.3% | 1.4% | 1.39% | [1.30%, 1.40%] | 900 |
| San Lazaro | 5.39% | 10.67% | 6.55% | [5.39%, 10.67%] | 100 |
| UP-NIH | 4.32% | 16.44% | 14.38% | [4.32%, 16.44%] | 80 |
| Lung Center | 8.51% | 9.85% | 10.02% | [8.51%, 10.02%] | 100 |
| BGHMC | 0.46% | 0.44% | 0.42% | [0.42%, 0.46%] | 150 |
| TMC | 2.39% | 2.75% | 5.88% | [2.39%, 5.88%] | 100* |
| St. Luke's, QC | 0.53% | 0.56% | 3.23% | [0.53%, 3.23%] | 100* |
| BPHL | 0.05% | 0.05% | 0.06% | [0.05%, 0.06%] | 100* |
| WVMC | 0.57% | 0.55% | 0.61% | [0.55%, 0.61%] | 80 |
| VSMC | 0.01% | 0.01% | 0.01% | [0.01%, 0.01%] | 240 |
| SPMC | 7.31% | 7.79% | 9.2% | [7.31%, 9.20%] | 100 |
| Caloocan City | 6.91% | 2.05% | 4.31% | [2.05%, 6.91%] | 100* |
| Cavite | 3.99% | 6.83% | 5.37% | [3.99%, 6.83%] | 100* |
| City of Mandaluyong | 6.33% | 3.59% | 4.64% | [3.59%, 6.33%] | 100* |
| City of Makati | 11.06% | 10.26% | 5.79% | [5.79%, 11.06%] | 100* |
| Laguna | 8.66% | 2.67% | 8.19% | [2.67%, 8.66%] | 100* |
| Taguig City | 8.72% | 9.04% | 8.23% | [8.23%, 9.04%] | 100* |
| Pasay City | 6.67% | 6.94% | 0.21% | [0.21%, 6.94%] | 100* |
| Rizal | 9.78% | 4.23% | 4.47% | [4.23%, 9.78%] | 100* |
| Bulacan | 4.81% | 1.64% | 3.44% | [1.64%, 4.81%] | 100* |
| City of Las Piñas | 2.23% | 2.23% | 3.61% | [2.23%, 3.61%] | 100* |

## Appendix B. Optimal solution for 30 testing centers

**Table B.3:** Proportion of test kits to be distributed in 30 testing centers.

| Testing Center | $k = 1.5614$ | $k = 0.4136$ | $k = 0.0849$ | Range | Daily limit |
| --- | --- | --- | --- | --- | --- |
| Research Institute for Tropical Medicine (RITM) | 1.32% | 1.21% | 1.51% | [1.21%, 1.51%] | 900 |
| San Lazaro Hospital | 2.81% | 6.99% | 3.51% | [2.81%, 6.99%] | 100 |
| UP-NIH | 5.41% | 6.76% | 7.19% | [5.41%, 7.19%] | 80 |
| Lung Center of the Philippines | 6.3% | 3.47% | 4.75% | [3.47%, 6.30%] | 100 |
| BGHMC | 0.43% | 0.43% | 0.43% | [0.43%, 0.43%] | 150 |
| TMC | 2.79% | 3.64% | 4.02% | [2.79%, 4.02%] | 100* |
| St. Luke's Medical Center, Quezon City | 0.37% | 0.41% | 1.1% | [0.37%, 1.10%] | 100* |
| BPHL | 0.05% | 0.05% | 0.06% | [0.05%, 0.06%] | 100* |
| WVMC | 0.57% | 0.61% | 0.62% | [0.57%, 0.62%] | 80 |
| VSMC | 0.01% | 0.01% | 0.01% | [0.01%, 0.01%] | 240 |
| SPMC | 2.41% | 2.4% | 5.27% | [2.40%, 5.27%] | 100 |
| Caloocan City | 4.9% | 6.12% | 3.02% | [3.02%, 6.12%] | 100* |
| Cavite | 3.32% | 3.2% | 3.38% | [3.20%, 3.38%] | 100* |
| City of Mandaluyong | 4.13% | 2.72% | 5.74% | [2.72%, 5.74%] | 100* |
| City of Makati | 6.56% | 3.09% | 1.74% | [1.74%, 6.56%] | 100* |
| Laguna | 4.65% | 2.86% | 5.32% | [2.86%, 5.32%] | 100* |
| Taguig City | 3.27% | 1.6% | 3.59% | [1.60%, 3.59%] | 100* |
| Pasay City | 2.93% | 5.01% | 0.3% | [0.30%, 5.01%] | 100* |
| Rizal | 4.01% | 3.91% | 3.67% | [3.67%, 4.01%] | 100* |
| Bulacan | 5.53% | 2.98% | 3.45% | [2.98%, 5.53%] | 100* |
| City of Las Piñas | 6.27% | 1.76% | 6.98% | [1.76%, 6.98%] | 100* |
| City of Parañaque | 2.02% | 6.07% | 4.74% | [2.02%, 6.07%] | 100* |
| City of Marikina | 4.50% | 4.96% | 4.51% | [4.50%, 4.96%] | 100* |
| City of Malabon | 2.11% | 3.33% | 1.22% | [1.22%, 3.33%] | 100* |
| City of Valenzuela | 3.51% | 3.51% | 3.56% | [3.51%, 3.56%] | 100* |
| City of Navotas | 7.06% | 8.68% | 4.67% | [4.67%, 8.68%] | 100* |
| Pangasinan | 5.72% | 5.31% | 3.42% | [3.42%, 5.72%] | 100* |
| Batangas | 3.27% | 3.17% | 4.94% | [3.17%, 4.94%] | 100* |
| Negros Occidental | 3.64% | 5.41% | 4.51% | [3.64%, 5.41%] | 100* |
| City of San Juan | 0.13% | 0.32% | 2.79% | [0.13%, 2.79%] | 100* |

